# A Genetic Perspective on Diabetic Complications: A Mendelian Randomization Study on the Increased Risk of Cataracts in Type 1 Diabetes

**DOI:** 10.1101/2024.01.29.24301964

**Authors:** Dongliang Pei, Shuyan Wang, Yuan Wang, Chenmin Sun

## Abstract

**Background:** The well-documented epidemiological link between type 1 diabetes and cataract is recognized. Yet, the question of a shared genetic foundation for these conditions, and the potential implication of a causal connection, is still an area of uncertainty.

**Methods:** We utilized a two-sample Mendelian randomization approach to extract and analyze four GWAS datasets from public databases, aiming to elucidate the causal relationship between Type 1 Diabetes and cataracts. Our study design incorporated multiple Mendelian randomization methods to assess the influence of Type 1 Diabetes genetic risk on the susceptibility to cataracts, and employed the MR-PRESSO global test for the evaluation of horizontal heterogeneity among the results.

**Results:** Mendelian Randomization analysis has indicated a statistically significant association between Type 1 Diabetes and an increased risk of cataract development. All applied Mendelian Randomization methods pointed to a positive causal estimate, suggesting that Type 1 Diabetes contributes to an elevated risk of cataracts.

**Conclusion:** This study supports the hypothesis of a causal relationship between Type 1 Diabetes and cataracts. Despite some heterogeneity, all outcomes are positively aligned, and leave-one-out analysis confirms the robustness of these findings. This research is significant for understanding the process of cataract prevention and treatment in patients with Type 1 Diabetes.

## Introduction

Type 1 Diabetes (T1D) represents a chronic condition that affects millions worldwide. The global incidence rate of T1D stands at 15 per 100,000 individuals, with a prevalence of 9.5%[1]. Notably, these rates are experiencing an upward trend[2, 3]. Epidemiological investigations have highlighted a significantly elevated risk of cataracts among patients with Diabetes, compared to the general populace[4, 5]. Most research on diabetes-induced cataracts has focused on metabolic mechanisms[6, 7], but often falls short in ruling out potential confounding factors, such as lifestyle elements and genetic predispositions [6]. The extent of genetic linkage between T1D and cataracts, and whether this linkage reflects a causal relationship, remains unclear.

Mendelian Randomization (MR), utilizing genetic variants as instrumental variables, emerges as a robust strategy for probing the potential causative link between diseases [8, 9]. This methodology circumvents the limitations commonly encountered in traditional observational studies, like confounding variables and reverse causation issues[10]. Our research leverages the two-sample MR approach, drawing on data from two separate Genome-Wide Association Studies (GWAS), to delve into the potential causal nexus between these two conditions. The strength of this approach lies in its use of genetic variations as instrumental variables, which remain unaffected by environmental factors post-birth, thus offering a distinctive lens to comprehend how T1D potentially escalates the risk of cataracts.

Through this investigation, our objective is to fortify the evidence base for the prevention and management of cataracts in individuals with diabetes, and to offer insights that could guide future research on biological mechanisms and public health intervention strategies.

## Materials and Methods

### Study Design

This study was conducted in strict adherence to institutional and national ethical guidelines. The data utilized were sourced from an open-source database, with original informed consent for the use of this data having been duly obtained by the respective GWAS teams. It is important to note that the data from the IEU platform consist of aggregate statistical summaries from GWAS, inherently devoid of any personally identifiable information, thereby ensuring ethical compliance in the use of this data.

This study utilizes the MR approach to investigate the potential causal link between T1D and cataract risk. We sourced our data from the IEU platform, an open-source database for GWAS summary data, established by the MRC Integrative Epidemiology Unit at the University of Bristol, UK[11, 12]. This analysis involved data from comprehensive European GWAS to examine the potential effects of diabetes-related genetic variants on cataract development risk. See Fig 1.

**Fig 1.**
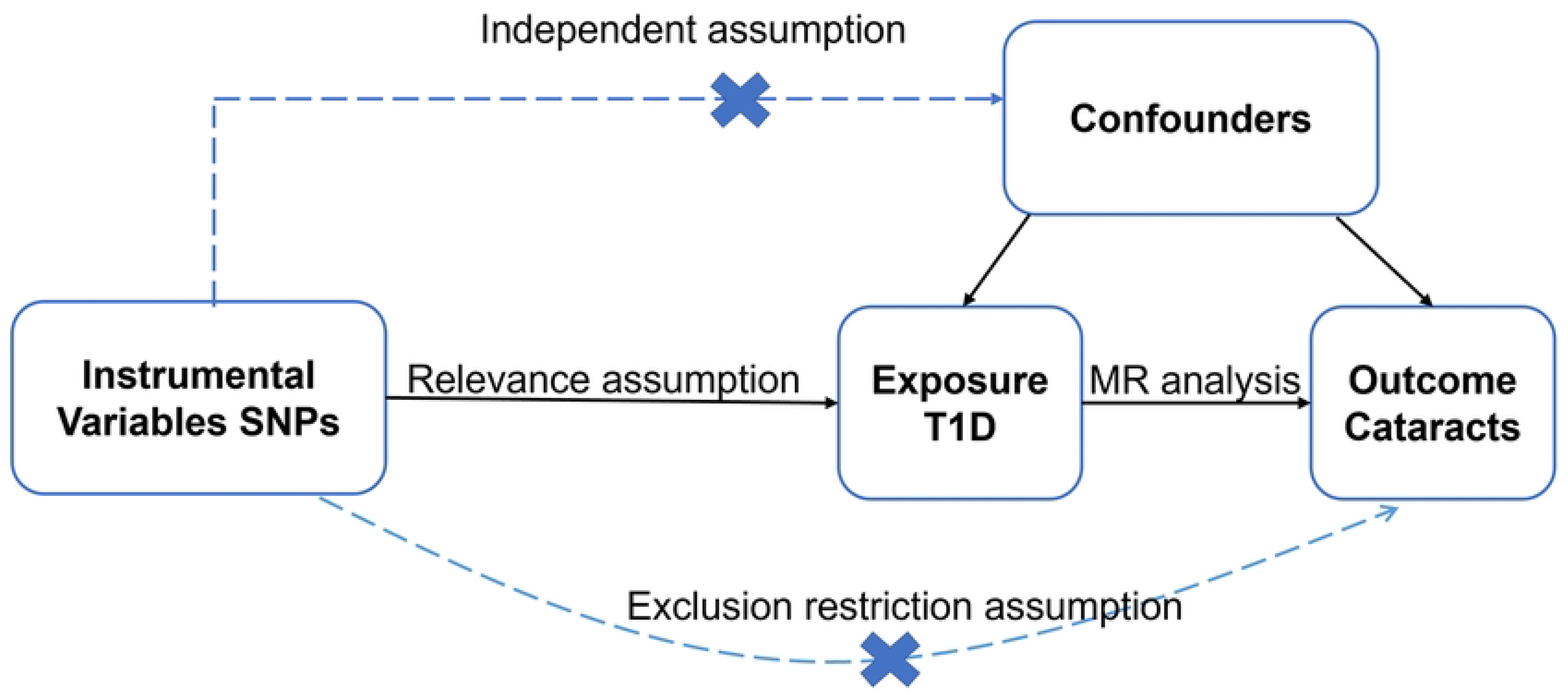
Design of the Mendelian Randomization Study of T1D and Cataracts. The instrumental variable in this Mendelian randomization study was selected based on the hypothesis that it is associated with T1D but not influenced by confounding variables. The hypothesis posits that this variable affects the risk of cataracts solely through its relationship with T1D.

### Data Source

Two independent sets of genetic instrumental variables associated with T1D from GWAS studies, ebi-a-GCST90018925 with a sample size of 457,695 individuals and 24,182,422 SNPs, and ebi-a-GCST90014023 with a sample size of 520,580 individuals and 59,999,551 SNPs, were extracted. We evaluated their impact on two cataract outcomes, ebi-a-GCST90018814 with a sample size of 491,877 individuals and 24,163,031 SNPs, and ebi-a-GCST90038649 with a sample size of 484,598 individuals and 9,587,836 SNPs[13-15], as detailed in Website: https://gwas.mrcieu.ac.uk/

### Data Integration and Processing

We employed the TwoSampleMR and MRPRESSO R packages for data integration and analysis. Relevant SNPs for diabetes (exposure) and cataract surgery (outcome) were extracted using extract_instruments and extract_outcome_data functions from the TwoSampleMR package. Data harmonization was achieved using harmonise_data, and linkage disequilibrium issues were managed with clump_data. The MR-PRESSO global test was used to address potential horizontal pleiotropy and outliers, including global outlier detection, correction, and distortion tests. SNPs related to outliers were excluded. Various MR methods, like MR Egger, weighted median, inverse-variance weighted, simple mode, and weighted mode, were utilized to explore the potential causal relationship between T1D and the need for cataract surgery. These methods aimed to quantify the effect of genetic predisposition to diabetes on the likelihood of requiring cataract surgery.

Heterogeneity was examined using the mr_heterogeneity function.

Leave-one-out analysis was conducted using the mr_leaveoneout function to assess the robustness of the results.

The robustness of the MR-PRESSO test results was evaluated using the mr_presso function.

#### Statistical Analysis

All analyses were performed using R software (version 4.3.2). The threshold for significance in MR analyses was set at P < 0.05. The same significance threshold was applied to outlier detection and distortion tests in the MR-PRESSO test.

## Results

We utilized extract_instruments and extract_outcome_data functions to extract relevant genetic instrumental variables and outcome data. In the data harmonization phase, linkage disequilibrium (LD) was addressed, and SNPs incompatible with the outcomes were excluded. Heterogeneity tests were also conducted to assess the consistency of the genetic instrumental variables in their impact on the outcomes.

MR analysis indicated a statistically significant association between T1D and two different cataract outcomes. In the analysis of the ebi-a-GCST90018814 outcome, using ebi-a-GCST90014023 as the exposure data, the inverse-variance weighted method suggested a causal association (b = 0.0132024389, p = 9.991590e-05), though the heterogeneity test showed marginal significant heterogeneity (Q_pval = 0.05240152). When using ebi-a-GCST90018925 as the exposure data, all MR methods pointed to a positive causal estimate, suggesting that T1D appears to increase the risk of cataracts. The p-values for almost all methods were extremely low, providing strong statistical evidence. The weighted median method showed the strongest causal association (b = 0.04585755, p = 1.785003e-07). See Fig 2. Table1.

**Fig 2.**
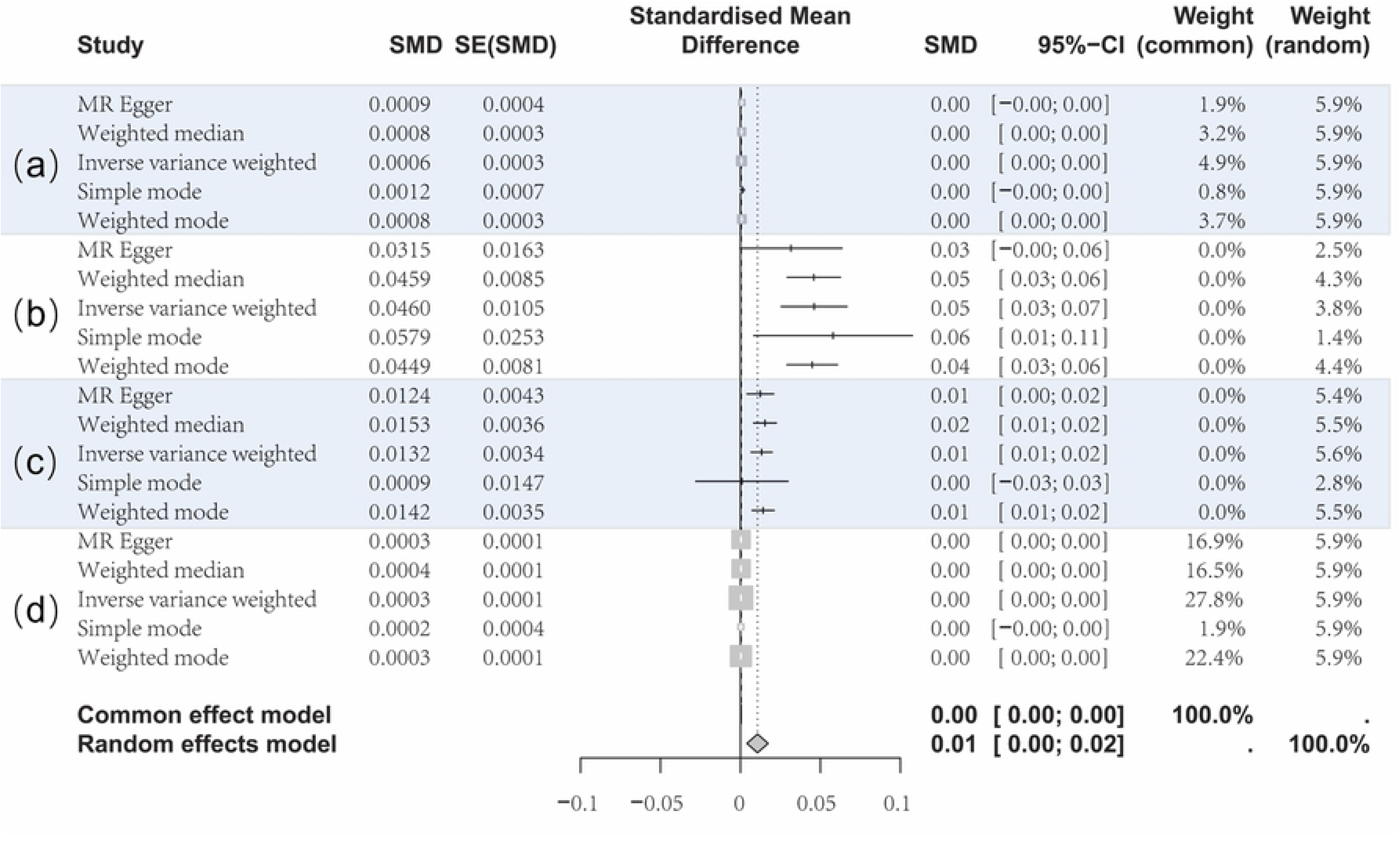
Forest Plot of Four Group Analysis of Two-Sample MR Methods. (a): ebi-a-GCST90018925 on ebi-a-GCST90038649, (b): ebi-a-GCST90014023 on ebi-a-GCST90018814, (c): ebi-a-GCST90014023 on ebi-a-GCST90018814, (d): ebi-a-GCST90014023 on ebi-a-GCST90018814. Despite the use of different MR methods potentially leading to varied effect sizes, the smaller standard errors and narrow confidence intervals signify the statistical validity of these estimates.

**Table 1.** Two-Sample MR Between Type 1 Diabetes and Cataracts.

| id.exposure | id.outcome | method | nsnp | b | se | pval |
| --- | --- | --- | --- | --- | --- | --- |
| ebi-a-GCST90018925 | ebi-a-GCST90038649 | MR Egger | 16 | 0.000867 | 0.000447 | 0.073076 |
|  |  | Weighted median | 16 | 0.000773 | 0.000343 | 0.024200 |
|  |  | IVW | 16 | 0.000605 | 0.000276 | 0.028573 |
|  |  | Simple mode | 16 | 0.001228 | 0.000677 | 0.089769* |
|  |  | Weighted mode | 16 | 0.000829 | 0.000318 | 0.019834 |
| ebi-a-GCST90018925 | ebi-a-GCST90018814 | MR Egger | 17 | 0.031537 | 0.016302 | 0.072145* |
|  |  | Weighted median | 17 | 0.045862 | 0.008516 | 7.22E-08 |
|  |  | IVW | 17 | 0.046036 | 0.010517 | 1.20E-05 |
|  |  | Simple mode | 17 | 0.057924 | 0.025322 | 0.036120 |
|  |  | Weighted mode | 17 | 0.044869 | 0.008073 | 4.33E-05 |
| ebi-a-GCST90014023 | ebi-a-GCST90018814 | MR Egger | 76 | 0.012421 | 0.004261 | 4.70E-03 |
|  |  | Weighted median | 76 | 0.015327 | 0.003646 | 2.62E-05 |
|  |  | IVW | 76 | 0.013202 | 0.003393 | 9.99E-05 |
|  |  | Simple mode | 76 | 0.000903 | 0.014722 | 0.951261* |
|  |  | Weighted mode | 76 | 0.014151 | 0.003528 | 1.41E-04 |
| ebi-a-GCST90014023 | ebi-a-GCST90038649 | MR Egger | 78 | 0.000309 | 0.000148 | 0.040659 |
|  |  | Weighted median | 78 | 0.000366 | 0.00015 | 0.014693 |
|  |  | IVW | 78 | 0.000305 | 0.000116 | 0.008325 |
|  |  | Simple mode | 78 | 0.00021 | 0.000438 | 0.632663* |
|  |  | Weighted mode | 78 | 0.000284 | 0.000129 | 0.030757 |

\* $pval > 0.05$

In the analysis of the ebi-a-GCST90038649 outcome, using both ebi-a-GCST90018925 and ebi-a-GCST90014023 as exposure data showed statistically significant associations, with the heterogeneity test showing no significant heterogeneity. This suggests more consistent effects of the genetic instrumental variables across these two datasets. See Fig 2. Table1.

### Heterogeneity Analysis

The heterogeneity test showed marginal significant heterogeneity (Q_pval = 0.05240152) in the analysis of the ebi-a-GCST90018814 outcome, using ebi-a-GCST90014023.The MR-PRESSO analysis did not identify any significant outliers, and the global test indicated no substantial data distortion. However, significant pleiotropy was detected in the analysis of the effect of ebi-a-GCST90018925 on ebi-a-GCST90018814, with a p-value of 0.021.

### Sensitivity Analysis

The leave-one-out sensitivity analysis further confirmed the robustness of our findings, with no single SNPs significantly altering the overall MR estimates, as shown in Fig 3.

**Fig 3.**
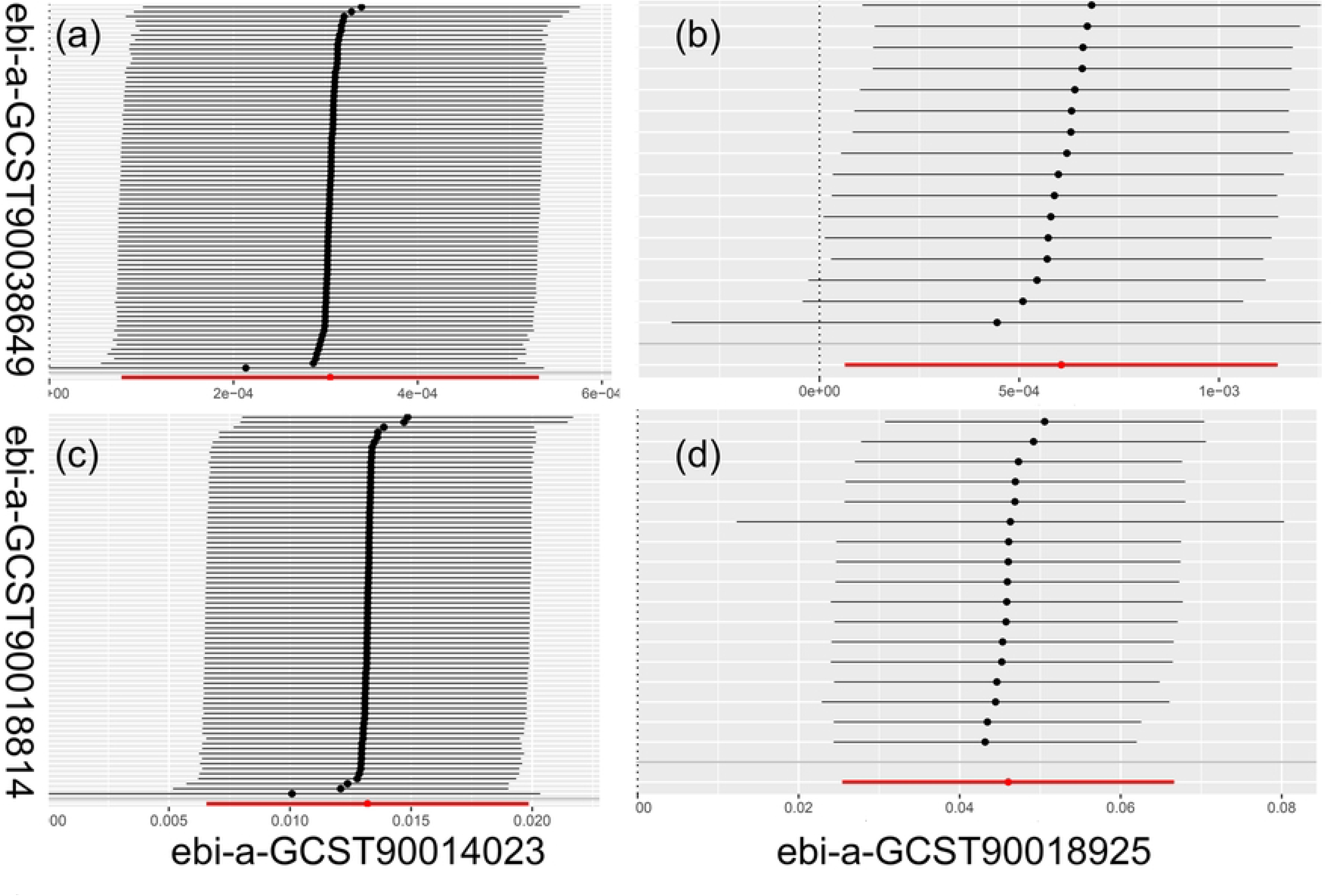
MR Leave-One-Out Sensitivity Analysis for the Impact of T1D on Cataracts. Each circle represents the MR analysis outcome for the remaining SNPs, after sequentially excluding each SNP in turn.

The scatter plots collectively demonstrate MR analyses using diverse MR methods, all indicating a consistent positive causal effect of T1D on the risk of developing cataracts. The uniform directionality of the SNP effect estimates across different analytical approaches reinforces the robustness of the causal inference. as shown in Fig 4.

**Fig 4.**
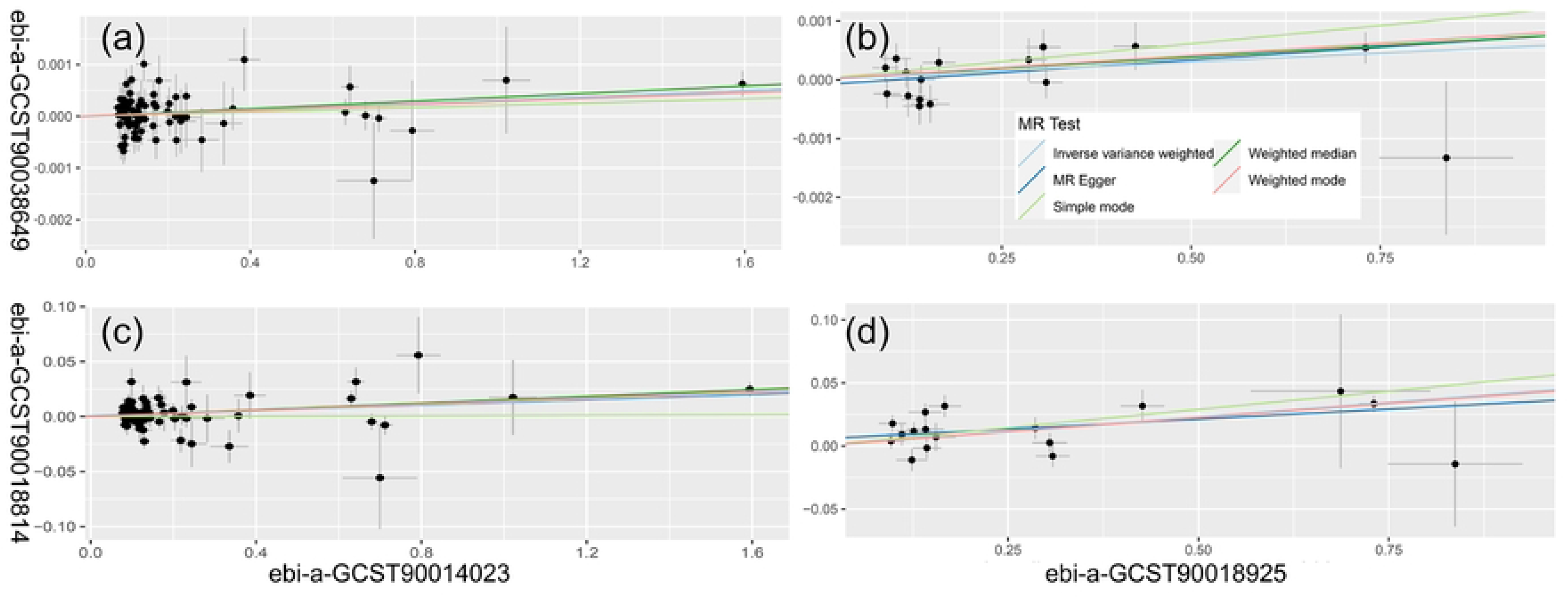
MR Analysis Scatter Plots for T1D and Cataract Risk. All indicating a consistent positive causal effect of T1D on the risk of developing cataracts. The uniform directionality of the SNP effect estimates across different analytical approaches reinforces the robustness of the causal inference.

## Discussion

This study employed a two-sample MR method to explore the potential causal relationship between T1D and cataracts. Our findings indicate a significant association between T1D and cataracts, suggesting a higher incidence of cataracts among diabetic patients. Although the MR-PRESSO global test detected significant pleiotropy in the effect of ebi-a-GCST90018925 on ebi-a-GCST90018814 (p-value: 0.021), the majority of our results are positive and robust, indicating that this does not compromise our main findings.

Diabetes is a metabolic disease where prolonged high blood sugar levels can lead to various complications. In ophthalmology, diabetes can affect the health of the lens in multiple ways, including accelerating cataract development through promoting non-enzymatic glycation reactions and oxidative stress[16-18]. It’s reported that the likelihood of cataracts in diabetic patients is five times higher. The risk is dependent on the duration of diabetes, the frequency of blood sugar levels exceeding the target range, and the presence of macular edema[19].

Patients with T1D have a higher risk of developing cataracts, which is a clouding of the eye’s lens leading to a decrease in vision and can affect the quality of life[19-21]. The occurrence of cataracts in diabetic patients is associated with prolonged uncontrolled high blood sugar and significant fluctuations in blood sugar levels, leading to osmotic imbalances in the lens. Risk factors for diabetic cataracts include elevated levels of glycated hemoglobin, diabetic ketoacidosis, and the duration and control of diabetes. Diabetic cataracts are more common in adolescent females and develop rapidly, often within weeks or months. The most common type of cataract in diabetic patients is posterior subcapsular cataracts[22].

MR is a research method that uses genetic variations as instrumental variables to examine the causal effects of modifiable exposures on diseases. Based on Mendel’s laws of inheritance and instrumental variable estimation, it can infer causal effects in the presence of unobserved confounders[8]. The term was initially proposed by Gray and Wheatley in 1991 and has been applied in studies using genetic variants reliably associated with modifiable exposures[23]. Compared to observational studies, this method is less likely to be affected by confounders or reverse causation[24]. MR has been applied in various fields, including medicine and public health, for causal inference. It has been used in studies concerning cancer, cardiovascular diseases, and more.

The study utilized Genome-Wide Association Study (GWAS) data from East Asian populations to conduct a MR analysis of Type 2 Diabetes and cataracts, integrating GWAS data with expression quantitative trait loci (eQTL) data. This identified two candidate functional genes (MIR4453HG and KCNK17), whose expression levels may be associated with the causal relationship between Type 2 Diabetes and cataracts[25]. our European study points to different genetic markers, indicating a complex and diverse genetic landscape influencing the link between diabetes and cataracts.

Despite the new insights our study offers into the relationship between T1D and cataracts, there are limitations. Firstly, our analysis relies on summary data from public GWAS databases, which may not capture all relevant genetic variations. Secondly, our analysis includes only data from European populations, limiting its generalizability.

Our findings have significant implications for public health strategies. Recognizing the higher risk of cataract development in T1D patients can promote earlier screening and intervention measures. Moreover, this discovery underscores the importance of blood sugar control in preventing cataracts in T1D patients.

## Conclusions

The results of this study support the hypothesis of a causal relationship between T1D and cataracts. Although there is heterogeneity, all outcomes are positive, and the leave-one plot indicates robust results. Particularly, the MR-PRESSO test results suggest that this relationship is unlikely to be affected by multidirectional heterogeneity. The directional consistency of the scatter plots is encouraging, and these findings are significant for understanding the process of preventing and treating cataracts in T1D patients.

## Data Availability

All relevant data are within the manuscript and its Supporting Information files.

## Acknowledgements

My heartfelt thanks go to my family and colleagues for their unwavering support and assistance. Your encouragement and cooperation have been invaluable on my journey.

## Supporting information

**S1_File.docx. ebi-a-GCST90018925 on ebi-a-GCST90038649**. the complete set of scripts and code used in the Mendelian Randomization study examining the effect of ebi-a-GCST90018925 on ebi-a-GCST90038649, facilitating replication and further exploration of the study’s methodology and findings.

**S2_File.docx. ebi-a-GCST90018925 on ebi-a-GCST90018814**. the complete set of scripts and code used in the Mendelian Randomization study examining the effect of ebi-a-GCST90018925 on ebi-a-GCST90018814, facilitating replication and further exploration of the study’s methodology and findings.

**S3_File.docx. ebi-a-GCST90014023 on ebi-a-GCST90018814**. the complete set of scripts and code used in the Mendelian Randomization study examining the effect of ebi-a-GCST90014023 on ebi-a-GCST90018814, facilitating replication and further exploration of the study’s methodology and findings.

**S4_File.docx. ebi-a-GCST90014023 on ebi-a-GCST90038649**. the complete set of scripts and code used in the Mendelian Randomization study examining the effect of ebi-a-GCST90014023 on ebi-a-GCST90038649, facilitating replication and further exploration of the study’s methodology and findings.

